# Knowledge and Attitude Towards Chronic Kidney Disease Among Residents in the Tamale Metropolis: A Cross-Sectional Study

**DOI:** 10.1101/2025.06.06.25329121

**Authors:** Abdul-Malik Abdulai, Samson Alhassan, Mohammed Saani Muftawu, Kabriku Peter Claver, Iddrisu Mohammed Sisala, Rashida Atrime, Abubakari Wuni, Mubarik Nngbanso Asumah

## Abstract

**Introduction:** Chronic Kidney Disease (CKD) affects 10-13% of the global population, with higher prevalence in regions like Sub-Saharan Africa and Ghana. It is linked to major non-communicable diseases and disproportionately impacts lower-middle-income countries. CKD often goes undiagnosed, with poor public awareness and misconceptions. This study aimed to assess the knowledge and attitudes of residents in Tamale Metropolis towards CKD.

**Method:** A quantitative cross-sectional study design was employed to collect data from 441 residents across eleven communities in the Tamale Metropolis. The data were analyzed using descriptive statistics, Chi-square tests, and logistic regression analysis.

**Results:** The overall knowledge of residents towards CKD was 54.0%, while 66.0% of them expressed a positive attitude toward CKD patients. Attitude toward CKD patients was significantly associated with gender, education level, occupation, employment status, and income (*p* < 0.001). Similarly, knowledge of CKD was significantly linked to gender, education level, occupation, and employment status (*p* < 0.001). The significant predictor of residents’ attitudes towards CKD was income levels (AOR = 3.39, *p* = 0.001). Also, significant predictors of knowledge on CKD were gender (AOR = 1.58, *p* = 0.04) and level of education (AOR = 7.16, *p* <0.01).

**Conclusion:** The results indicate general knowledge of CKD, yet significant gaps remain in knowledge about kidney function and risk factors. Additionally, while income status and gender were significant predictors of knowledge, education had a substantial impact on attitudes toward CKD. These results underscore the need for targeted educational initiatives to reduce stigma and enhance understanding across diverse demographics.

## Background

Chronic kidney disease is a major silent killer globally, making it a significant public health burden in both developed and developing countries [1, 2]. CKD is defined as decreased glomerular filtration rate, increased excretion of urinary albumin, or both. [3]. The global prevalence has been projected to be 10-13% [4], and that of sub-Saharan Africa was revealed to be 13.9 % [5–7]. In Ghana, the prevalence of chronic renal disease is 13.3 % [8].

Globally, approximately 850 million people are reported to have kidney disease, with a majority living in lower-middle-income countries (LMICs), where resource constraints limit access to diagnosis, prevention, or treatment of the disease [4]. Furthermore, data show that as many as nine out of ten individuals living with kidney disease in these resource-poor settings are unaware that they have the condition and therefore do not seek treatment [9, 10].

Given the chronic yet symptomatic nature of CKD, it is important that not only healthcare providers but also ordinary people are knowledgeable and have a favourable attitude towards its diagnosis and treatment [11]. However, numerous studies from multiple countries have reported poor knowledge and attitudes towards CKD among various at-risk populations. In a study by Yusoff, Yusof and Kueh [12] in Malaysia, most respondents had poor knowledge but demonstrated good attitudes and practices toward CKD risks. In Palestine, Sa’adeh, Darwazeh [13] reported that age and educational attainment were significantly associated with higher CKD knowledge scores among respondents. A Saudi Arabian study revealed that CKD knowledge was correlated with age, educational attainment, monthly income, civil status, physical activity habits, and medical history [14]. In Jordan, Khalil and Abdalrahim [15] reported that although most participants were knowledgeable about kidney disease, half held incorrect information regarding CKD signs and symptoms. In Africa, a study on CKD in northern Tanzania revealed that community-based adults had limited knowledge of CKD, with an average knowledge score of 3.28 out of 10. Despite this, there was a modest foundation upon which to build awareness programs. Concerning the attitude towards CKD, a study in Indonesia by Agustiyowati [16] revealed a high positive attitude towards CKD prevention and control was observed among the educated population The study further indicated that educational level significantly impacts patients’ attitudes towards kidney disease prevention. Yusoff, Yusof and Kueh [12] also identified occupation, marital status, sex, and age as key predictors of a favourable attitude toward CKD. Other factors such as age, monthly income, and a high knowledge score have also been shown to be strongly associated with better CKD attitude scores [15]. However, Stanifer, Turner [17] reported that people’s attitudes towards CKD treatment can be affected by concerns about the health, economic, and social impacts of the disease, or hope for recovery and diet [16].

Kidney research in Ghana, however, has largely focused on health workers’ knowledge and care practices, with little attention paid to the knowledge and attitudes of the general populace towards the condition [1]. Research findings from a Ghanaian study examining healthcare-seeking behaviour among CKD patients in a tertiary facility in northern region found that participants had varying levels of knowledge about their condition, which significantly influenced their health-seeking behaviours [18]. Generally, participants demonstrated positive attitudes and behaviours towards seeking healthcare, relying primarily on hospital services and expressing trust in healthcare providers. However, nearly all participants faced financial difficulties in affording CKD-related treatments. Only a few considered seeking herbal treatments in addition to hospital care. This cross-sectional study is conceptualized to assess the knowledge and attitude of residents of Tamale Metropolis on chronic kidney disease. Specifically, this study assessed the knowledge of residents on chronic kidney disease, determined the attitudes of residents toward CKD and examined the influence of socio-demographics of residents on knowledge and attitude about CKD in the Tamale Metropolis.

## Methods and Materials

### Study design

This study utilized a descriptive cross-sectional design with a quantitative approach to collect data from residents regarding CKD.

### Study setting

The study was conducted in the Tamale Metropolis, located in the Northern Region of Ghana, established by Legislative Instrument [L.I. 2068]. As the regional and metropolitan capital, Tamale is strategically positioned with a land size of 646.9 km² and serves as a market hub for goods from neighbouring regions and countries like Burkina Faso, Niger, Mali, and Togo. The metropolis comprises 116 communities, with 35% urban, 13% peri-urban, and 52% rural, where agriculture plays a key role, though these rural areas face challenges in infrastructure such as roads, schools, and healthcare facilities.

### Study population

The study population, comprising residents of the Tamale Metropolis, was drawn from a total of 418,520 individuals as reported in the District Health Information Management System 2 (DHIMS2) data [19]. This depicts the area’s significance as a growing urban center in Northern Ghana.

### Inclusion and exclusion criteria

This study included residents of the Tamale Metropolis who had lived in the area for at least six months, were 18 years or older, and had no serious physical or mental illness, provided they were willing to participate. Respondents who did not provide voluntary consent were excluded from the study.

### Sample size determination

Yamane [20] simplified sample size formula was employed in determining the sample size for this study. According to Yamane’s formula, given a desired level of precision, desired confidence level and the estimated proportion of the attribute present in the population, the sample size was calculated as follows:

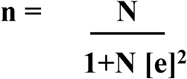

where n is the sample size, N is the total population of Tamale Metro (418,520) [19], and e is the margin of error [0.05].

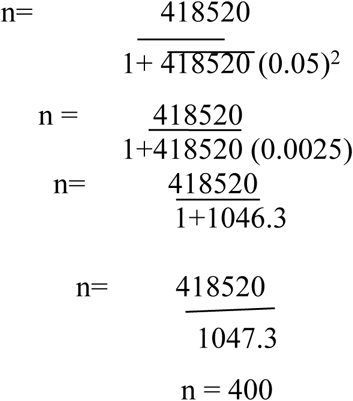

This figure was increased by 10% to make room for non-response, increased participation and possible bias. Thus, the total sample size for this study was 440. However, at the end of the data collection, 441 responses were obtained and utilised for the final analysis.

### Sampling techniques

Cluster simple random sampling was used to select the participants. The metropolis has four sub-districts; each sub-district was treated as a separate cluster. Applying the probability proportional to size approach, the sample allocation for the four (4) sub-districts was estimated. Simple random sampling was then used to select the desired number of participants in each sub-district for the study. This approach reduced survey costs and time by narrowing the sample in stages, making data collection more manageable while ensuring a diverse and representative sample.

### Data collection instrument

The study adopted portions of the questionnaires developed by Alobaidi [14] on knowledge about CKD with a Cronbach’s coefficient value of 0.71 and [21] on attitudes toward individuals living with chronic diseases with a Cronbach’s coefficient value of 0.88. The complete tool for this study consisted of three sections. Section A gathered socio-demographic information, while Section B assessed knowledge of CKD with 24 multiple-choice items using the options ‘True’, ‘False’, and ‘I don’t know’. Section C measured attitudes toward CKD with 10 items using a 4-point Likert scale: strongly disagree = 1, disagree = 2, agree = 3, and strongly agree = 4. This subscale was slightly modified in wording to suit the context of the study, with adjustments mainly made for clarity, such as the use of chronic kidney disease and other terminology.

### Validity and reliability

The instrument was pretested among 30 residents of Doyinayili using the same inclusion and exclusion criteria. Following the pretest, items that were unclear or confusing were reworded. The revised instrument was then reviewed by a panel of three experts in the field to ensure content validity before the actual data collection began. The knowledge section on chronic kidney disease had a Cronbach’s alpha coefficient of 0.67 while the attitudes section, measuring attitudes toward chronic kidney disease, demonstrated good internal consistency with a Cronbach’s alpha coefficient of 0.81.

### Data collection procedure

Data collection commenced after obtaining ethical approval. In addition to the authors, 15 research assistants were trained in ethical considerations, data collection procedures, Kobo Collect tool and communication skills under the supervision of the principal investigator. The Trained research assistants, fluent in both English and Dagbani were sent to the selected communities and households to administer the questionnaire. During the administration phase, the data collection team used a Kobo Collect tool to administer the questionnaire. This made the process more efficient and allowed for quicker response collection and was specifically designed to minimize the likelihood of respondents skipping questions. To maintain ethical standards, informed consent was obtained from all respondents. The collected data was anonymized and securely stored to ensure respondent privacy.

### Data analysis

The data was exported to Microsoft Excel (version 19) for review and completeness checks, then cleaned and transferred to Stata software (version 17) for analysis. Descriptive analysis summarized socio-demographics, knowledge, and attitudes using frequencies and percentages in tables. Inferential analysis, including Chi-square tests and logistic regression, examined the influence of socio-demographic factors on knowledge and attitudes. Statistical significance was set at a p-value of less than 0.05.

### Ethical considerations

The research complied with principles outlined in the Declarations of Helsinki and sought ethical approval from the Human Research, Publications & Ethics Committee of Kwame Nkrumah University of Science and Technology with reference number CHRPE/AP/990/24 Permission was obtained from the Tamale Metropolitan Assembly (TMA) and community leaders, including assemblymen and chiefs, to conduct the study. Respondents provided written and oral consent after receiving the necessary information. Participation was voluntary, and individuals could withdraw at any time. The study findings would be shared with the public through open-access academic journals, the institutional website and repository, social media, and presentations at public lectures, webinars, and conferences.

### Results Respondent’s socio-demographic information

A total of 441 residents participated in this study, representing a response rate of more than 100%. Among them, the majority (57.8%) were aged between 30 and 59 years. Men accounted for 58.0% of the respondents, and 60.0% were married. In terms of education, most residents (34.7%) had completed tertiary education. Regarding occupation, 32.4% were primarily engaged in farming or trading, while 31.1% worked in skilled or technical professions, either within the formal sector or through various informal jobs. Furthermore, 65.3% of residents were employed full-time or part-time in the formal sector. Most residents (64.8%) earned an average monthly income of less than 1,000 Ghana cedis (see Table 1).

**Table 1:**
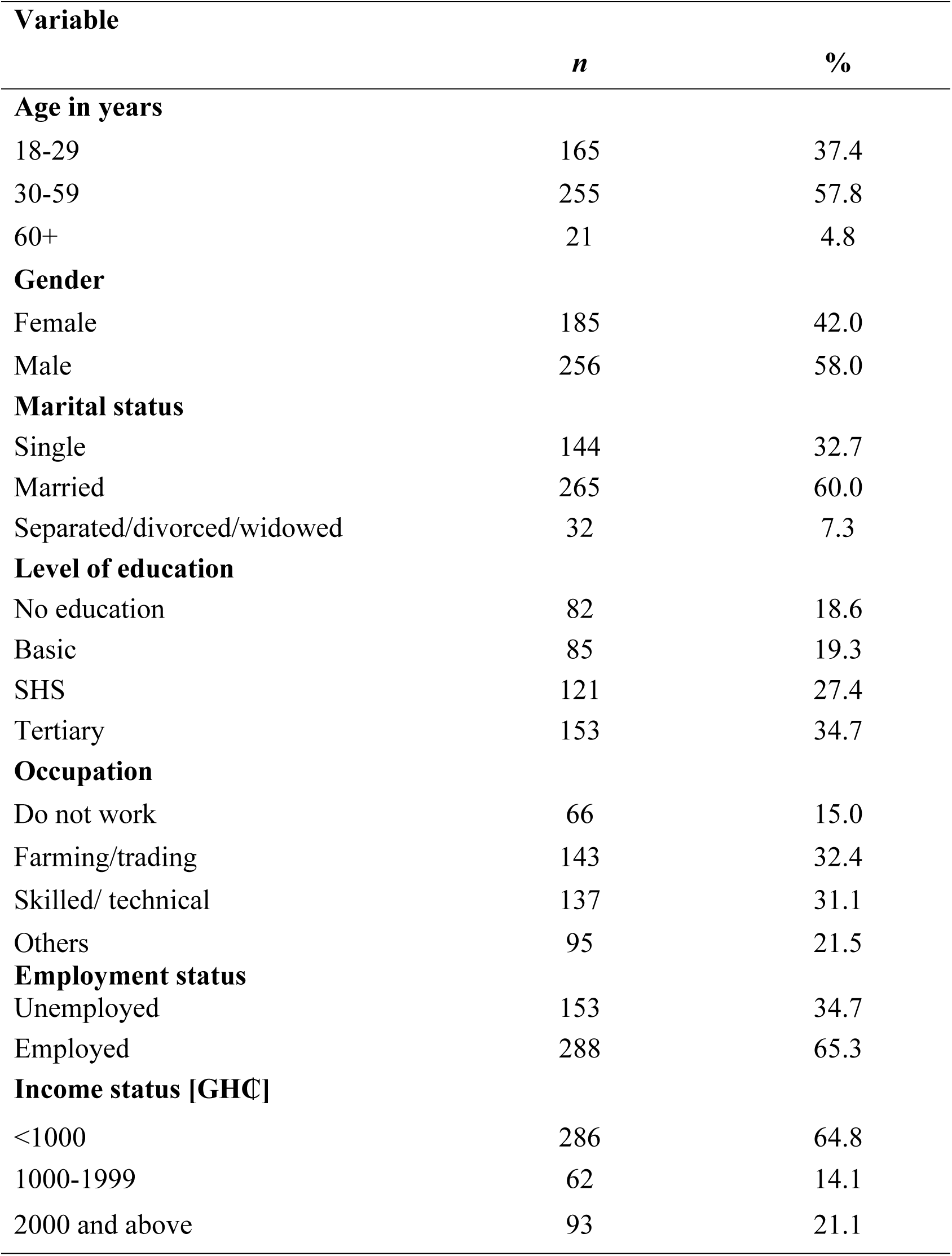
Respondent’s socio-demographic information.

### Knowledge of residents towards chronic kidney disease

In this study, the overall level of knowledge among residents regarding CKD was found to be 54.0%. When examining specific areas of knowledge, 53.6% of residents demonstrated an understanding of the signs and symptoms associated with CKD. In relation to diagnostic procedures, 58.4% of residents recognized the necessity of hospital tests and acknowledged that urine characteristics could be instrumental in the diagnostic process. Furthermore, 70.6% of residents were aware that an individual could live with a single healthy kidney and understood the roles of medications, dialysis, and professional guidance in the management and prevention of CKD. However, residents exhibited a lack of sufficient knowledge concerning kidney functions, with only 48.2% demonstrating awareness in this area. Furthermore, a limited number (39%) of residents correctly identified key risk factors for CKD, including diabetes, hypertension, obesity, female gender, excessive stress, and heart attack. These findings are detailed in Table 2.

**Table 2:** Knowledge of residents towards CKD.

| Knowledge | *False/I don't know* |  | True |  |
| --- | --- | --- | --- | --- |
|  | n | % | n | % |
| **Functions of the kidney** |  |  |  | **48.2** |
| The kidneys make urine. | 202 | 45.8 | 239 | 54.2 |
| The kidneys filter waste products from the blood. | 181 | 41.0 | 260 | 59.0 |
| The kidneys clean blood. | 176 | 39.9 | 265 | 60.1 |
| The kidneys help to keep blood sugar levels normal. | 252 | 57.6 | 187 | 42.4 |
| The kidneys help to maintain blood pressure | 255 | 57.8 | 186 | 42.2 |
| The kidneys help to break down protein in the body. | 271 | 61.4 | 170 | 38.6 |
| The kidneys help to keep the bones healthy. | 262 | 59.4 | 179 | 40.6 |
| **Risk factors for CKD** |  |  |  | **39.0** |
| Diabetes | 217 | 49.2 | 224 | 50.8 |
| Being a female gender | 357 | 81.0 | 84 | 19.0 |
| High blood pressure | 257 | 58.3 | 184 | 41.7 |
| Heart problems such as heart failure or heart attack | 240 | 54.4 | 201 | 45.6 |
| Excess stress | 260 | 59.0 | 181 | 41.0 |
| Obesity | 280 | 63.5 | 161 | 36.5 |
| **Signs and symptoms of CKD** |  |  |  | **53.6** |
| Water retention [excess water in the body] | 201 | 45.6 | 240 | 54.4 |
| Fever. | 202 | 45.8 | 239 | 54.2 |
| Nausea and vomiting | 282 | 64.0 | 159 | 36.0 |
| Loss of appetite | 278 | 40.4 | 263 | 59.6 |
| Increased fatigue [tiredness] | 159 | 36.0 | 282 | 64.0 |
| **Investigations** |  |  |  | **58.4** |
| A person can tell if he/she has kidney disease just by the colour, quality, or smell of urine | 285 | 64.6 | 156 | 35.4 |
| Kidney disease can only be diagnosed by a test at the hospital. | 82 | 18.6 | 359 | 81.4 |
| **Other considerations** |  |  |  | **70.6** |
| A person can lead a normal life with one healthy kidney. | 171 | 38.8 | 270 | 61.2 |
| Certain medications can help to slow-down the worsening of chronic kidney disease. | 137 | 31.0 | 304 | 69.0 |
| Dialysis is a form of treatment for kidney disease. | 152 | 34.5 | 289 | 65.5 |
| Kidney disease can be prevented if you follow the advice of a health professional. | 59 | 13.4 | 382 | 86.6 |
| **Overall knowledge of CKD** |  |  |  | **54.0** |

### Attitude towards CKD

The study assessed residents’ attitudes toward CKD. Overall, 66.0% of residents have a positive attitude towards CKD patients. However, less than half (48.3%) agreed that people with CKD are as intelligent as others, and 49.7% believed there should be restrictions for those with the condition. However, a majority (59.9%) felt that individuals with CKD should be approached with caution, yet most (70.5%) affirmed the disease does not define a person, and 69.8% believed they are not different from others. A significant proportion (66.4%) did not view the disease as a result of weakness or foolishness, and 69.6% agreed that those with CKD could safely work with children. Conversely, only a small percentage (19.7%) thought individuals with CKD should feel ashamed, and 22.4% believed they should be segregated in the workplace. Details are presented in Table 3.

**Table 3:** Attitude of residents towards CKD.

|  | *Strongly disagree/*<br>*Disagree* |  | *Strongly agree/*<br>*agree* |  |
| --- | --- | --- | --- | --- |
|  | n | % | n | % |
| **Attitude** |  |  |  | **66.0** |
| People with chronic kidney diseases are as intelligent as anybody else | 228 | 51.7 | 213 | 48.3 |
| People with chronic kidney diseases should be approached with caution. | 177 | 40.1 | 264 | 59.9 |
| Having chronic kidney disease doesn't define who you are. | 130 | 29.5 | 311 | 70.5 |
| People with chronic kidney diseases are not different from others | 133 | 30.2 | 308 | 69.8 |
| People with chronic kidney diseases should not be ashamed of themselves | 87 | 19.7 | 354 | 80.3 |
| People with chronic kidney diseases have no reason to feel guilty | 144 | 32.7 | 297 | 67.4 |
| Chronic kidney diseases are not a result of being weak-willed or foolish | 148 | 33.6 | 293 | 66.4 |
| People with chronic kidney diseases should not be segregated at the workplace | 99 | 22.4 | 342 | 77.6 |
| People with chronic kidney diseases can safely work with children | 134 | 30.4 | 307 | 69.6 |
| People with chronic diseases must not expect some restrictions on their freedom | 219 | 49.7 | 222 | 50.3 |

### Association between socio-demographics of residents and attitude

The association between demographic characteristics of residents and attitudes was assessed using a Chi-square (χ²) test. The analysis indicated statistically significant associations between gender, education level, occupation, employment status, income status and attitude (*p* < 0.05). In contrast, no statistically significant associations were found between age, marital status, and attitude (*p* > 0.05) (Table 4).

**Table 4:** Association between socio-demographics of residents and attitude.

| Variable | ATTITUDE | | | | $\chi^2$ | *p*- value |
| --- | --- | --- | --- | --- | --- | --- |
|  | Unfavourable attitude |  | Favourable attitude |  |  |  |
|  | *n* | % | *n* | % |  |  |
| **Age [yrs]** |  |  |  |  |  |  |
| 18-29 | 65 | 39.4 | 100 | 60.6 | 0.548 | 0.76 |
| 30-59 | 101 | 39.6 | 154 | 60.4 |  |  |
| 60+ | 10 | 47.6 | 11 | 52.4 |  |  |
| **Gender** |  |  |  |  |  |  |
| Female | 86 | 46.5 | 99 | 53.5 | 5.749 | **0.02** |
| Male | 90 | 35.2 | 166 | 64.8 |  |  |
| **Marital status** |  |  |  |  |  |  |
| Single | 53 | 36.8 | 91 | 63.2 | 1.305 | 0.52 |
| married | 108 | 40.8 | 157 | 59.2 |  |  |
| Separated/divorced/widowed | 15 | 46.9 | 17 | 53.1 |  |  |
| **Level of education** |  |  |  |  |  |  |
| No education | 39 | 47.6 | 43 | 52.4 | 24.810 | **0.001** |
| Basic | 44 | 51.8 | 41 | 48.2 |  |  |
| SHS | 56 | 46.3 | 65 | 53.7 |  |  |
| Tertiary | 37 | 24.2 | 116 | 75.8 |  |  |
| **Occupation** |  |  |  |  |  |  |
| Do not work | 24 | 36.4 | 42 | 63.6 | 12.470 | **0.006** |
| Farming/trading | 73 | 51.0 | 70 | 49.0 |  |  |
| Skilled/ technical | 51 | 37.2 | 86 | 62.8 |  |  |
| Others | 28 | 29.5 | 67 | 70.5 |  |  |
| **Employment** |  |  |  |  |  |  |
| Unemployed | 43 | 28.1 | 110 | 71.9 | 13.613 | **0.001** |
| Employed | 133 | 46.2 | 155 | 53.8 |  |  |
| **Income status [GHC]** |  |  |  |  |  |  |
| <1000 | 137 | 47.9 | 149 | 52.1 | 27.814 | **0.001** |
| 1000-1999 | 23 | 37.1 | 39 | 62.9 |  |  |
| 2000 and above | 16 | 17.2 | 77 | 82.8 |  |  |
| **\*$p < 0.05$, \*\*$p < 0.001$** |  |  |  |  |  |  |

### Association between socio-demographics of residents and knowledge

The association between respondents’ demographic characteristics and knowledge was analyzed using a chi-square (χ²) test. The findings revealed statistically significant associations between gender, education level, occupation, employment status, and knowledge (*p* < 0.01). However, no statistically significant associations were observed between age, marital status, income level, and knowledge (*p* > 0.05), as presented in Table 5.

**Table 5:** Association between socio-demographics of residents and knowledge.

| Variable | KNOWLEDGE | | | | $\chi^2$ | *p*-value |
| --- | --- | --- | --- | --- | --- | --- |
|  | Inadequate knowledge |  | Knowledge |  |  |  |
|  | *n* | % | *n* | % |  |  |
| Age [yrs] |  |  |  |  |  |  |
| 18-29 | 69 | 41.8 | 96 | 58.2 | 3.097 | 0.21 |
| 30-59 | 110 | 43.1 | 145 | 56.9 |  |  |
| 60+ | 13 | 61.9 | 8 | 38.1 |  |  |
| Gender |  |  |  |  |  |  |
| Female | 101 | 54.6 | 84 | 45.4 | 15.850 | 0.001 |
| Male | 91 | 35.6 | 165 | 64.4 |  |  |
| Marital status |  |  |  |  |  |  |
| Single | 56 | 38.9 | 88 | 61.1 | 2.138 | 0.34 |
| married | 120 | 45.3 | 145 | 54.7 |  |  |
| Separated/divorced/widowed | 16 | 50.0 | 16 | 50.0 |  |  |
| Level of education |  |  |  |  |  |  |
| No education | 56 | 68.3 | 26 | 31.7 | 65.634 | **0.001** |
| Basic | 51 | 60.0 | 34 | 40.0 |  |  |
| SHS | 55 | 45.5 | 66 | 54.5 |  |  |
| Tertiary | 30 | 19.6 | 123 | 80.4 |  |  |
| **Occupation** |  |  |  |  |  |  |
| Do not work | 27 | 40.9 | 39 | 59.1 | 12.463 | **0.006** |
| Farming/trading | 79 | 55.2 | 64 | 44.8 |  |  |
| Skilled/ technical | 53 | 38.7 | 84 | 61.3 |  |  |
| Others | 33 | 34.7 | 62 | 65.3 |  |  |
| **Employment** |  |  |  |  |  |  |
| Unemployed | 45 | 29.4 | 108 | 70.6 | 19.017 | **0.001** |
| Employed | 147 | 51.0 | 141 | 49.0 |  |  |
| **Income status [GHC]** |  |  |  |  |  |  |
| <1000 | 132 | 46.2 | 154 | 53.8 | 5.000 | 0.08 |
| 1000-1999 | 29 | 46.8 | 33 | 53.2 |  |  |
| 2000 and above | 31 | 33.3 | 62 | 66.7 |  |  |

**\* $p < 0.05$ , \*\* $p < 0.001$**

### Influence of socio-demographic characteristics on attitude of residents

A logistic regression analysis was performed to assess the impact of residents’ socio-demographic characteristics on attitudes. The results indicated that income status (AOR = 3.39, *p* = 0.001) was a significant predictor of attitude. In contrast, gender, education level, occupation, and employment status did not demonstrate any statistically significant effects on attitude (Table 6).

**Table 6:** Influence of socio-demographics of residents on attitude.

| **Model** | **Variable** | **AOR** | ***p*-value** | **95% C.I.** |
| --- | --- | --- | --- | --- |
|  | **Gender** |  |  |  |
|  | Female | Ref | 0.48 |  |
|  | Male | 1.17 |  | 0.759 – 1.792 |
|  | **Level of education** |  |  |  |
|  | No education | Ref |  |  |
|  | Basic | 0.8 | 0.40 | 0.403 – 1.438 |
|  | SHS | 0.97 | 0.93 | 0.539 – 1.753 |
|  | Tertiary | 1.62 | 0.13 | 0.861 – 3.056 |
|  | **Occupation** |  |  |  |
|  | Do not work | Ref |  |  |
|  | Farming/trading | 0.54 | 0.06 | 0.285 – 1.021 |
|  | Skilled/ technical | 0.78 | 0.45 | 0.407 – 1.483 |
|  | Others | 0.94 | 0.86 | 0.461 – 1.906 |
|  | **Employment** |  |  |  |
|  | Unemployed | Ref |  |  |
|  | Employed | 0.83 | 0.45 | 0.504 – 1.353 |
|  | **Income status (GHC)** |  |  |  |
|  | <1000 | Ref |  |  |
|  | 1000-1999 | 1.52 | 0.17 | 0.840 – 2.750 |
|  | 2000 and above | 3.39 | **0.001** | 1.791 – 6.418 |

*\*p* < .05, *\*\*p* < .001

### Influence of socio-demographics of residents on knowledge

Logistic regression analysis was used to determine if the socio-demographic information of residents would have a significant influence on knowledge. The results showed that gender (AOR = 1.58, *p* < 0.05) and level of education (AOR = 7.16, *p* <0.01) were found to be significant predictors of knowledge. Nonetheless, the rest of the socio-demographic information of residents statistically had no predictive effect on knowledge. Details are summarized in Table 7.

**Table 7:** Influence of socio-demographics of residents on knowledge.

| **Model** | **Variable** | **AOR** | ***p*-value** | **95% C.I.** |
| --- | --- | --- | --- | --- |
|  | **Gender** |  |  |  |
|  | Female | Ref |  |  |
|  | Male | 1.58 | **0.04** | 1.019 – 2.446 |
|  | **Level of education** |  |  |  |
|  | No education | Ref |  |  |
|  | Basic | 1.39 | 0.33 | 0.721 – 2.680 |
|  | SHS | 2.19 | **0.01** | 1.188 – 4.022 |
|  | Tertiary | 7.16 | **0.001** | 3.602 – 14.239 |
|  | **Occupation** |  |  |  |
|  | Do not work | Ref |  |  |
|  | Farming/trading | 0.93 | 0.82 | 0.473 – 1.812 |
|  | Skilled/ technical | 1.09 | 0.80 | 0.551 – 2.167 |
|  | Others | 0.95 | 0.90 | 0.455 – 2.003 |
|  | **Employment** |  |  |  |
|  | Unemployed | Ref |  |  |
|  | Employed | 0.67 | 0.13 | 0.403 – 1.117 |
|  | **Income status [GHC]** |  |  |  |
|  | <1000 | Ref |  |  |
|  | 1000-1999 | 0.66 | 0.18 | 0.354 – 1.221 |
|  | 2000 and above | 0.72 | 0.29 | 0.386 – 1.326 |

**\**p* < .05, \*\**p* < .001**

## Discussion

This study assessed residents’ knowledge and attitudes toward chronic kidney disease in the Tamale Metropolis. It also examined the influence of the socio-demographics of residents on knowledge and attitude about CKD.

In this study, residents demonstrated an overall knowledge of CKD with an average score of 54.0%. Specific areas of strength included a recognition of the importance of hospital tests and urine characteristics in the diagnosis and monitoring of CKD (58.4%), an understanding that individuals can live with a single healthy kidney (70.6%), and awareness of the roles of medications, dialysis, and professional guidance in the management and prevention of CKD (70.6%). These findings suggest that while residents may need improvement in their overall knowledge of CKD, their understanding of its management, diagnostic tools, and kidney function is more robust. This finding is consistent with the results of studies conducted in Indonesia by Calisanie, Isyaturodhiyah and Lindayani [22] and in Lebanon by Younes, Mourad [23], both of which reported that residents demonstrated a sufficient level of knowledge regarding CKD. Therefore, healthcare professionals need to strengthen health education initiatives to improve public knowledge of CKD.

However, 53.6% of residents exhibited an understanding of the signs and symptoms associated with CKD, while 48.2% demonstrated insufficient knowledge regarding kidney function. Furthermore, a limited proportion of residents (39%) accurately identified key risk factors for CKD, such as diabetes, hypertension, obesity, female gender, excessive stress, and heart attack. These findings suggest that there are notable gaps in residents’ understanding of both the physiological aspects of kidney health and the factors that contribute to the development of CKD, highlighting areas where further education and training are needed. This study’s finding contrasts with the results of other studies in which more than half of the respondents correctly identified diabetes mellitus (DM), hypertension (HTN), and obesity as risk factors for CKD [14, 24]. Similarly, a Malaysian study by Yusoff, Yusof and Kueh [12] also reported poor knowledge of CKD among participants. Participants’ general lack of knowledge of the risk factors for CKD could explain why the disease is becoming increasingly common among persons in developing countries like Ghana [8]. Moreover, an Indonesian study by Calisanie, Isyaturodhiyah and Lindayani [22] reported that half of the respondents had incorrect information about the symptoms of CKD. This could indicate a gap in public knowledge or understanding of the disease, which could potentially affect people’s ability to recognize CKD symptoms early and seek proper medical help. This also suggests that there may be a need for better education and awareness programs related to CKD, as people might be misinformed or unaware of its signs and symptoms.

In this study, it was reported that majority (66%) of residents exhibited a positive attitude toward individuals with CKD. Despite the overall positive outlook, certain misconceptions and biases remain, highlighting the need for further attention and intervention. Most participants demonstrated positive perceptions, with 70.5% affirming that CKD does not define a person. Similarly, 69.8% believed individuals with CKD are not different from others, and 67.4% supported the view that those affected should not feel guilty about their condition. This reflects a growing awareness that CKD is a medical condition rather than a moral or personal failure. These findings align with results from a study conducted by Yusoff, Yusof and Kueh [12], where over 65% of participants viewed CKD as a manageable health condition, and stigmatization was less prevalent in urban populations. Additionally, 69.6% of participants agreed that individuals with CKD could safely work with children, underscoring the recognition that CKD does not compromise one’s competence or safety. These attitudes align with evidence suggesting that individuals with CKD can maintain normal societal roles with adequate management [16]. Also, the present study found that 66.4% of respondents did not consider CKD a result of being weak-willed or foolish. In contrast to this finding Gebreyohannes, Wake and Abdulle [25] reported that 45% of rural participants in Sub-Saharan Africa believed CKD was caused by supernatural forces or poor life choices. The observed differences may be ascribed to superior education and increased access to information in urban centers such as Tamale, which likely mitigate the impact of detrimental cultural beliefs. In contrast, the tendency of most residents to exercise caution in their interactions with CKD patients underscores a significant area of concern. These misconceptions emphasize the need for targeted educational interventions to address and dispel these unfounded fears.

Despite these positive findings, some participants exhibited discriminatory attitudes. For instance, 49.7% believed restrictions should be imposed on individuals with CKD, and 59.9% felt they should be approached with caution. These perceptions suggest lingering fears and misconceptions about the disease. Similarly, 19.7% of participants agreed that individuals with CKD should feel ashamed, and 22.4% supported workplace segregation. Such stigmatizing attitudes could impede social integration and mental well-being for those living with CKD. A similar trend was observed in a study by Rashid, Deshwal [26], where 20% of participants believed CKD was contagious, leading to unnecessary precautions and exclusion in workplace settings.

This study explored the impact of residents’ socio-demographic factors on their knowledge of CKD. The results indicated that gender was significantly correlated with CKD knowledge, with males demonstrating a higher level of awareness compared to females. This finding is consistent with a survey conducted in Malaysia, which similarly reported that females exhibited poorer knowledge of CKD than males [12]. The disparity in CKD knowledge between males and females has been documented in previous research [11]. Thus, the limited knowledge observed among female participants in this present study may be attributed to their socio-demographic background, as majority were not employed or were housewives, and had limited formal education.

The educational level of respondents emerged as a significant determinant of their knowledge regarding CKD. The results of this study revealed a positive correlation, indicating that individuals with higher educational attainment demonstrated greater knowledge of CKD. This finding aligns with the research conducted by Mahmoud, Ibrahim [27] who identified higher education as a significant predictor of increased knowledge of CKD. In contrast, other studies have reported significantly lower CKD knowledge scores among respondents [28, 29]. Despite the presence of contrary findings, higher education remains a key determinant, as it enhances access to health information, improves health literacy, and strengthens individuals’ capacity to understand and apply knowledge related to CKD. Therefore, healthcare authorities within urban areas should prioritize the expansion of educational initiatives on kidney diseases to further enhance the knowledge base of the population.

This study found that the socio-demographic factor of income significantly influenced individuals’ attitudes toward CKD. Specifically, higher income status was a significant predictor of more positive attitudes toward CKD, suggesting that individuals with higher income levels are more likely to exhibit favourable attitudes toward the disease. This finding aligns with the results of previous studies, which suggest that individuals’ attitudes toward concerns about kidney disease and its social implications are predominantly shaped by more favourable economic conditions [11, 30]. This implies that improved economic conditions can enhance individuals’ ability to access quality healthcare, engage in educational initiatives, and utilize mental health resources and social support networks, thereby fostering proactive attitudes toward the prevention and management of diseases such as CKD.

### Implications and recommendations

This study reveals the key findings regarding knowledge and attitudes toward CKD in the Tamale Metropolis. While respondents demonstrated a basic understanding of kidney functions, significant gaps remained in their awareness of more complex functions like blood sugar regulation, blood pressure maintenance, and bone health. Knowledge of CKD risk factors, such as high blood pressure, heart disease, and obesity, was also insufficient, along with limited understanding of diagnostic tools like urine testing. Despite these gaps, there was a positive outlook on kidney health maintenance, particularly regarding medications and dialysis. The study also revealed a need for public health campaigns to address misconceptions and reduce stigma, particularly concerning discriminatory attitudes in social and workplace settings. Gender, education, and income disparities were observed, with males, individuals with higher education, and higher-income groups demonstrating better knowledge and more positive attitudes toward CKD. To improve awareness and attitudes, targeted public health campaigns should focus on education tailored to diverse groups, engage community leaders, and work to reduce health-related stigma, particularly in lower-income communities.

## Conclusion

The findings indicate that while there is general knowledge of CKD, significant gaps remain in knowledge regarding kidney function and risk factors. Furthermore, although 66% of residents exhibit positive attitudes toward individuals with CKD, further efforts are needed to reduce stigma, promote inclusion, and educate the public to better support individuals living with the condition.

The study also found that residents’ income status was a significant predictor of their attitude toward CKD, with higher income levels correlating with more positive attitudes, emphasizing the importance of addressing socioeconomic disparities in shaping public perceptions of CKD. Furthermore, gender and educational level were key predictors of knowledge about CKD, with males and individuals with higher education levels demonstrating greater knowledge. These results underscore the need for targeted, inclusive, and multifaceted educational initiatives that address gender differences and varying educational backgrounds to improve public knowledge and attitudes toward CKD.

## Acknowledgements

The authors would like to first express their sincere gratitude to the Tamale Metropolitan Assembly and the Assembly Members of the various communities for granting permission and providing support during the data collection process. Secondly, we extend our special thanks to the respondents, whose participation in this study is deeply appreciated. Finally, the authors are deeply grateful to the final-year nursing students of NMTC, Tamale, for their invaluable assistance in the data collection.

## Data availability

The data used in this study are available and can be provided upon reasonable request from the corresponding author.

## Authors’ Contribution

Conceptualization: AA; Data collection: AS, MMS, PKC, SM; Formal analysis: AS, MMS; Manuscript original draft: AS, MMS, PKC, SMI, A; Manuscript review and editing: AA, AS, MMS, AMN, AW

